# Specific epigenetic age acceleration measures are associated with oral health outcomes in U.S. adults

**DOI:** 10.64898/2026.06.16.26354138

**Authors:** Andrea L. Tang, Amy Tsurumi

## Abstract

**Objectives:** Oral health conditions impact a significant proportion of the global population. Chronological age is a known risk factor; however, characterization of epigenetic age remains limited and is expected to provide additional insight into biological mechanisms.

**Materials and Methods:** The National Health and Nutrition Examination Survey (NHANES) was used to analyze the effect of epigenetic age measures of DunedinPoAm, and epigenetic age acceleration (EAA) of Horvath, Hannum, Weidner, Lin, VidalBralo, PhenoAge, GrimAge, and GrimAge2, on various oral health outcomes from survey and examination results. Univariable and multivariable logistic regression were performed, adjusting for sex, race-ethnicity, education, poverty income ratio categories, and dental insurance coverage status.

**Results:** DunedinPoAm was associated with the last dental appointment being for an existing issue (p=0.0093), poor general oral condition (p=0.0226), limiting food due to teeth problems (p=0.0031), and recommendation to see a dentist within the next two weeks (p=0.0171). EAAs for PhenoAge, GrimAge, and GrimAge2, were associated with a smaller number of oral health outcomes, whereas EAAs for Horvath, Hannum, Weidner, Lin, and Vidal-Bralo showed no associations.

**Conclusions:** In a representative U.S. population, DunedinPoAm was most consistently positively associated with different adverse oral health outcomes compared with other epigenetic aging measures.

**Clinical Relevance:** Tracking specific epigenetic ages such as DunedinPoAm, EAA GrimAge, EAA GrimAge2, and PhenoAge, may aid in additional monitoring of oral health outcomes. Understanding specific aging-related CpGs associated with oral health may aid in elucidating underlying molecular mechanisms.

## Introduction

Oral health conditions are estimated to impact approximately 3.69 billion globally, and given its public health significance, the WHO Global Oral Health Action Plan has called for efforts towards a 10% reduction in their prevalence by 2030,^1–4^ highlighting the importance of characterizing risk factors. Aging is a major risk factor for oral diseases, with numerous studies having demonstrated mechanistic links.^5–7^ While most studies focused on the impact of chronological age, analysis of molecular markers of aging, such as 5-methylcytosine DNA methylation (DNAm), are limited and expected to provide additional insight into biological mechanisms.

Data from the National Health and Nutrition Examination Survey (NHANES), which was recently released for the first time, enables a population-based study of epigenetic aging and oral health outcomes. While recent studies analyzing the NHANES epigenetic age data have reported links with cardiovascular disease and all-cause mortality,^8,9^ mortality among pre-diabetes and diabetes subpopulations,^10^ and those with cardiovascular-kidney-metabolic syndrome,^11^ hypertension,^12^ coronary heart disease,^13^ socioeconomic status,^14^ race and ethnicity,^15^ physical activity,^16^ and urinary heavy metal levels,^17^ one investigating oral health outcomes is lacking. Previous NHANES studies have shown associations between periodontitis and biological age measures of homeostatic dysregulation (HD),^18,19^ Klemera-Doubal method (KDM),^20^ and PhenoAge^21^ (based on blood chemistries only),^22^ or with KDM and PhenoAge, and further showed their impact on mortality^23^ or cognitive function.^24^ Another NHANES study showed a link between periodontitis and telomere length.^25^ Taken together, a study examining oral health variables in relation to epigenetic aging acceleration is missing.

Given this gap in prior literature, the goal of this study was to evaluate the association between oral health outcomes and epigenetic aging, among the U.S. general population. Major epigenetic clocks in the NHANES include Horvath,^26^ Hannum,^27^ Weidner,^28^ Lin,^29^ and VidalBralo,^30^ developed to predict chronological age. Others developed considering clinical variables, include DunedinPoAm^31^ (46 CpGs predicting the pace of aging, developed from 18 markers related to oral health, cardiovascular, respiratory, kidney, liver, body composition, and metabolic and immune functions with the Dunedin Study birth cohort^32^), PhenoAge^33^ (513 CpGs predicting phenotypic age based on chronological age and nine blood biomarkers for liver, kidney, liver, metabolic, and immune functions), and GrimAge^34^ and GrimAge2^35^ (panels of predictors from 1030 CpG sites for smoking pack-years and various blood protein assessments, with age and sex, to predict time-to-mortality).

Comparing whether specific epigenetic aging clocks are more strongly associated with oral health outcomes than others, and whether they are more predictive than chronological age, is expected to inform the development of additional strategies for monitoring oral health. Notably, DunedinPoAm included oral (gum) health among the pace of life markers for its development, and therefore, we hypothesized that it may be more strongly associated with oral health outcomes than other DNAm clocks.

## Methods

### Study Population

A cross-sectional study was conducted using publicly available data from the National Health and Nutrition Examination Survey (NHANES), an annual survey of a representative sample of the US civilian, noninstitutionalized population conducted by the Centers for Disease Control and Prevention (CDC).^36^ A non-human subjects research determination was obtained to analyze this data. The STROBE checklist for cross-sectional studies was used to guide the reporting of the results.

## Variables used

NHANES 1999-2000 and 2001-2002 included several epigenetic age clock data based on Illumina EPIC 850K DNA methylome array data (released later on July 31, 2024).^37^ We used DunedinPoAm,^31^ developed to predict the pace of aging, Horvath,^26^ Hannum,^27^ Weidner,^28^ Lin,^29^ and VidalBralo,^30^ developed to predict chronological age, PhenoAge,^33^ developed to predict phenotypic age, and GrimAge^34^ and GrimAge2,^35^ developed to predict time-to-mortality. Variables used for demographic and socioeconomic status data (original variable names and survey indications from NHANES are provided in parentheses) were: survey data on sex (RIAGENDR: male, female), age at screening in years (RIDAGEYR) or exam age in months (RIDAGEEX), race/ethnicity (RIDRETH2: “non-Hispanic White, non-Hispanic Black, Mexican American, other race including multi-racial, other Hispanic), poverty income ratio (PIR) (INDFMPIR), educational level (DMDEDUC2: less than 9^th^ grade, 9-11 grade, high school graduate/GED or equivalent, some college or AA degree, college graduate or above), and whether insurance covered any part of dental care (HID040: yes, no). Due to smaller numbers, the “other race including multi-racial” and “other Hispanic” categories were combined as “Other.” PIR was categorized into “low” for 0 to <1.3, “middle” for ≥1.3 to <4, and “high” for ≥4. Educational level of less than 9^th^ grade and 9-11 grade were combined as “not high school graduate.”

Oral health outcomes were based on survey data questions about the main reason for the last dentist appointment (OHQ033), general oral condition of the mouth and teeth (OHQ010), and whether participants limit foods because of teeth problems (OHQ020), and examination results data on overall recommendation (OHAREC), untreated caries/restorative needs (OHAROCDT), periodontal needs (OHAROCGP), and gingival needs (OHAROCOH). These survey or examination data were used to classify participants as having oral health issues or not, as shown in Supplementary Table S1.

## Inclusion and exclusion criteria

Since this study entailed analysis of existing survey data, the study size was determined by the inclusion/exclusion criteria. Among the 21,004 participants in the 1999-2000 and 2001-2002 data combined, a total of 721 were analyzed (flowchart of included/excluded numbers provided in Supplementary Figure S1).

The following participants were excluded: DNAm age data unavailable or XY chromosome predictions not matching reported sex; age at screening (RIDAGEYR) or examination (RIDAGEEX) reported as 85 (NHANES data recodes all ages 85 or above as 85 for confidentiality); missing data or reported “refused” or “don’t know” for PIR (INDFMPIR), education status (DMDEDUC2), or dental insurance coverage (HID040); missing data for the last dentist appointment (OHQ030) or indicating more than three years; oral health examination (OHAEXSTS), dentition examination (OHASCST3), or referral status (OHASCST5) reported as “partial” or “not done”; oral health examination screening questionnaire answers (OHQ130, OHQ132, OHQ134, OHQ136, OHQ138, OHQ140, OHQ142, OHQ144, OHQ146, OHQ148) reported as “yes”; examined edentulous status (OHXEDEN) indicated as “yes”; none of the 32 teeth examined (OHX01TC to 32TC) reported as “permanent” (*i.e.* all indicated as “implant” or “not present” or “missing”); dental outcomes of interest (OHQ033, OHQ010, OHQ020, OHAREC, OHAROCDT, OHAROCGP, OHAROCOH) reported as “don’t know.”

## Statistical Analysis

R (version 4.4.2)^38^ was used. To account for the complex, multistage, stratified, clustered probability sampling design, survey-weighted analyses were conducted using the “WTDN4YR” weights from the DNAm dataset, with the “survey (version 4.4.2)”^39–41^ package. Age at examination, recorded in months, was converted to age for the analysis. Epigenetic Age Acceleration (EAA) was obtained as the residual from linear regression of each DNAm age (independent variable) on chronological age (dependent variable), as is conventionally conducted.^34,42,43^ Correlations between different ages were conducted by weight-adjusted linear regression with summary statistics obtained with the “jtools (version 2.3.1)”^44^ package. To evaluate baseline characteristics and compare between groups, continuous variables were summarized as mean ± SD, with two-sample p-values. For categorical variables, unweighted counts, weighted percentage, and weighted adjusted Wald^45^ p-values were calculated. Quasibinomial logistic regression was performed as a univariable model with each epigenetic age measure as a predictor, or as a multivariable model adjusting for sex, race-ethnicity, PIR categories, educational status, and dental insurance status. Plots were constructed using the “ggplot2 (version 4.0.2)”^46^ and “gridExtra (version 2.3)”^47^ packages. Comparisons between the <50 and ≥50 age groups at examination were made with the overall 1999-2000 and 2001-2002 combined data (regardless of DNAm and oral health data), using the “WTMEC4YR” weights from the demographics data table.

## Results

### Baseline characteristics

The overall baseline characteristics are shown in Table 1, with weighted mean ± standard deviation (SD) or unweighted counts and weighted proportions. Chronological age was 61.57 ± 9.31, with 53.0% female and 47.0% male. The largest race-ethnicity category was non-Hispanic White, with 83.7%. The relatively large female and non-Hispanic White proportions are consistent with age ≥ 50 group (53.5% female and 79.3% non-Hispanic White), as compared to the < 50 group (50.7% female and 68.0% non-Hispanic White), for the overall population (regardless of having DNAm or oral health outcomes reported) (Supplementary Table S2), indicating a shift in the demographics for the older U.S. population. The largest PIR categories were middle-high (≥1.3 to 4) with 37.2% and high (≥4) with 54.5%. The largest education categories were college graduate and above with 33.3% and high school graduate with 27.5%. The proportion of participants with insurance partially or fully covering dental care was 58.3%. The baseline DunedinPoAm was 1.08 ± 0.09, and other epigenetic age acceleration (EAA) measures ranged from -0.08 ± 6.39 for EAA GrimAge2 to 0.21 ± 5.63 for EAA VidalBralo.

**Table 1.** Baseline characteristics. Survey-weighted mean ± standard deviation for continuous variables, and unweighted count (weighted %) for categorical variables are shown. EAA: epigenetic age acceleration.

|  | Overall (N=721) |
| --- | --- |
| **Chronological age** | 61.6 ± 9.3 |
| **Sex:** |  |
| Female | 343 (53.0%) |
| Male | 378 (47.0%) |
| **Race-Ethnicity:** |  |
| Mexican American | 160 (2.5%) |
| Non-Hispanic Black | 142 (6.9%) |
| Non-Hispanic White | 358 (83.7%) |
| Other Hispanic and Other including multi-racial | 61 (6.9%) |
| **PIR category:** |  |
| Low (<1.3) | 109 (8.4%) |
| Middle-high (≥1.3 to 4) | 306 (37.2%) |
| High (≥4) | 306 (54.5%) |
| **Education:** |  |
| Not high school graduate | 188 (13.7%) |
| High school graduate | 175 (27.5%) |
| Some college of AA graduate | 168 (25.5%) |
| College graduate or above | 190 (33.3%) |
| **Insurance includes dental coverage:** |  |
| Yes | 443 (59.3%) |
| No | 278 (40.7%) |
| **Epigenetic age:** |  |
| DunedinPoAm | 1.08 ± 0.09 |
| EAA Horvath | 0.06 ± 6.88 |
| EAA Hannum | 0.07 ± 6.89 |
| EAA Weidner | 0.11 ± 10.20 |
| EAA Lin | 0.12 ± 9.46 |
| EAA VidalBralo | 0.21 ± 5.63 |
| EAA PhenoAge | 0.11 ± 8.04 |
| EAA GrimAge | -0.06 ± 5.88 |
| EAA GrimAge2 | -0.08 ± 6.39 |

## DNAm clocks are correlated with chronological age, whereas DunedinPoAm and EAA are not

As expected, association with chronological age was not found for DunedinPoAm, which measures the pace of life (R^2^=0.002, p=0.501), whereas they were for Horvath, Hannum, Weidner, Lin, VidalBralo, PhenoAge, GrimAge, and GrimAge2 (R^2^ ranging from 0.289 to 0.698, p<0.00001 for all) (Supplementary Figure S2A). It is common to evaluate EAAs instead to remove confounding by chronological age, and as expected, the EAAs were no longer associated with chronological age (R^2^ ranging from 0.00001 to 0.00046, p-value ranging from 0.643 to 0.961). Another approach is to assess the gap (*i.e.* difference) between chronological and DNAm ages; however, they are known to be systematically smaller at higher chronological ages, and a negative correlation was found for most of the gap measures in this dataset as well (R^2^ ranging from 0.029 to 0.602; p<0.001 for all). Thus, further analysis was performed using EAA, as it is expected to be the most effective. EAA for PhenoAge, GrimAge, and GrimAge2, which are developed for phenotypic age or mortality, were significantly correlated with DunedinPoAm, whereas those based on chronological age prediction were not (Horvath, Hannum, Weidner, Lin, and VidalBralo) (Supplementary Figure S2B).

## Specific DNAm clocks are higher among those with adverse oral health outcomes

To test our overall hypothesis that adverse oral health outcomes may be associated with higher DunedinPoAm or various EAA measures, we comprehensively evaluated seven outcomes recorded in both surveys (1. whether the main reason for the last dentist appointment was due to an existing oral issue or not for any existing issue; 2. general oral condition self-reported as being poor versus very good, good, or fair; and 3. reported limiting food intake due to teeth problems as always, very often, often, or sometimes versus seldom or never), and examination results (1. overall recommendations to see a dentist within two weeks versus seeing one at the earliest convenience or continuing with regular routine care; 2. reporting untreated caries or restorative needs; 3. reporting periodontal needs; and 4. reporting gingival needs) (Supplementary Table S1).

First, we evaluated whether the mean DunedinPoAm or EAAs were significantly higher among those with adverse oral health outcomes than without. While chronological age did not show significant increase, except for the overall examination recommendation (p=0.003), DunedinPoAm was significantly higher for the last dentist appointment being for an existing issue (p=0.008), general oral condition being poor (p=0.019), limiting food intake due to teeth problems (p=0.002), and recommendation to see a dentist within two weeks (p=0.035) (Table 2). On the other hand, EAAs for Horvath, Hannum, Weidner, Lin, and VidalBralo were not significantly different for any of the seven outcomes, while the EAA PhenoAge was higher for poor general condition (p=0.041), limiting food (p=0.035), and overall recommendation to see a dentist within two weeks (p=0.011), and the EAA GrimAge and GrimAge2 were higher for the last dentist appointment being for an existing issue (p=0.006 and p=0.003, respectively), poor general oral condition (p=0.009 and p=0.011, respectively), and periodontal needs (p=0.009 and p=0.011, respectively) (Supplementary Table S3).

**Table 2.**
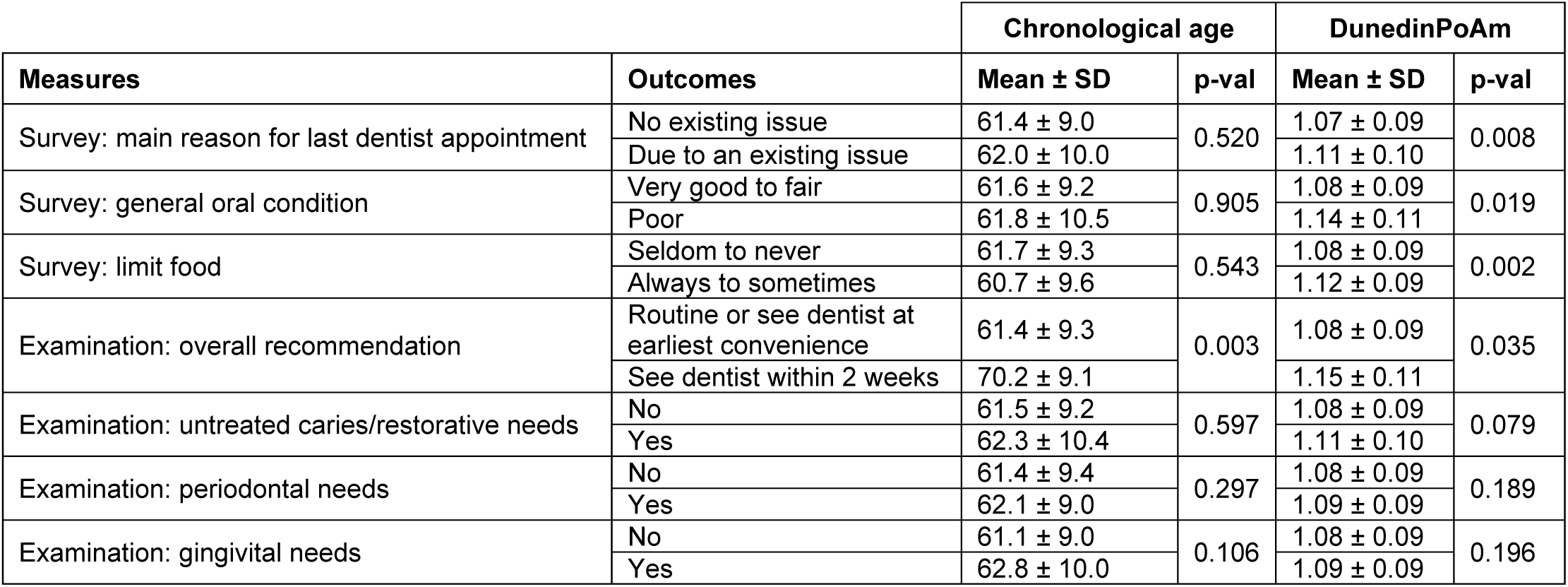
Differences in the means of chronological age and DunedinPoAm of the oral health outcomes. Survey-weighted mean ± standard deviation and two-sample t-test p-values are shown.

For demographic and socioeconomic status, the proportions of sex were not notably different for any of the oral health outcomes, while the relative proportion of Non-Hispanic White was lower, and other race-ethnicity categories were higher among those limiting food due to teeth problems (p=0.023), and examination indicating restorative (p=0.001) and gingival need (p=0.016) (Supplementary Table S4). A relatively lower proportion of the high PIR category and higher proportion of middle and low categories were found for poor general oral condition (p=0.015), limiting food due to teeth problems (p=0.042), recommendation to see a dentist within two weeks (p=0.042), and restorative (p=0.030), periodontal (p=0.009), and gingival needs (p=0.043). A relatively higher proportion of less than high school or high school graduate, and a lower proportion of college or associate degree, or higher groups were found for the main reason for the last dentist appointment due to an existing issue (p=0.015), and restorative (p=0.004), and gingival needs (p=0.019). A higher proportion of those without dental insurance coverage was found only among those reporting poor general oral condition (p=0.029).

## Univariable and multivariable models show positive associations between specific DNAm ages and oral health outcomes

Univariable and multivariable models (adjusted for sex, race-ethnicity, PIR categories, education categories, and dental insurance status) were constructed to assess the impact of different age measures on the seven adverse oral health outcomes (Figure 1A-B; Supplementary Table S5, S6). Chronological age was only associated with recommendation to see a dentist within the next two weeks (univariable p=0.0036, multivariable p=0.0053) (Supplementary Table S5, S6). DunedinPoAm, on the other hand, was positively associated with the last dental appointment being for an existing issue (p=0.0034), poor general oral condition (p=0.0083), limiting food due to teeth problems (p=0.0010), recommendation to see a dentist within the next two weeks (p=0.0097), and restorative needs (p=0.0493) in the univariable model (Figure 1A). These associations remained significant in the multivariable model for the last dental appointment being for an existing issue (p=0.0093), poor general oral condition (p=0.0226), limiting food due to teeth problems (p=0.0031), and recommendation to see a dentist within the next two weeks (p=0.0171) (Figure 1B). In these multivariable models, restorative need was positively associated with Non-Hispanic Black (p<0.0001) and Other (p=0.0349), while negatively associated with the low PIR category (p=0.0348), and college graduate or above, compared to no high school diploma (p=0.0060). Periodontal need was negatively associated with high PIR (p=0.0320), and gingival need was positively associated with the Non-Hispanic Black race-ethnicity category (p=0.0050) and negatively associated with college graduate or above (p=0.0243).

**Figure 1.**
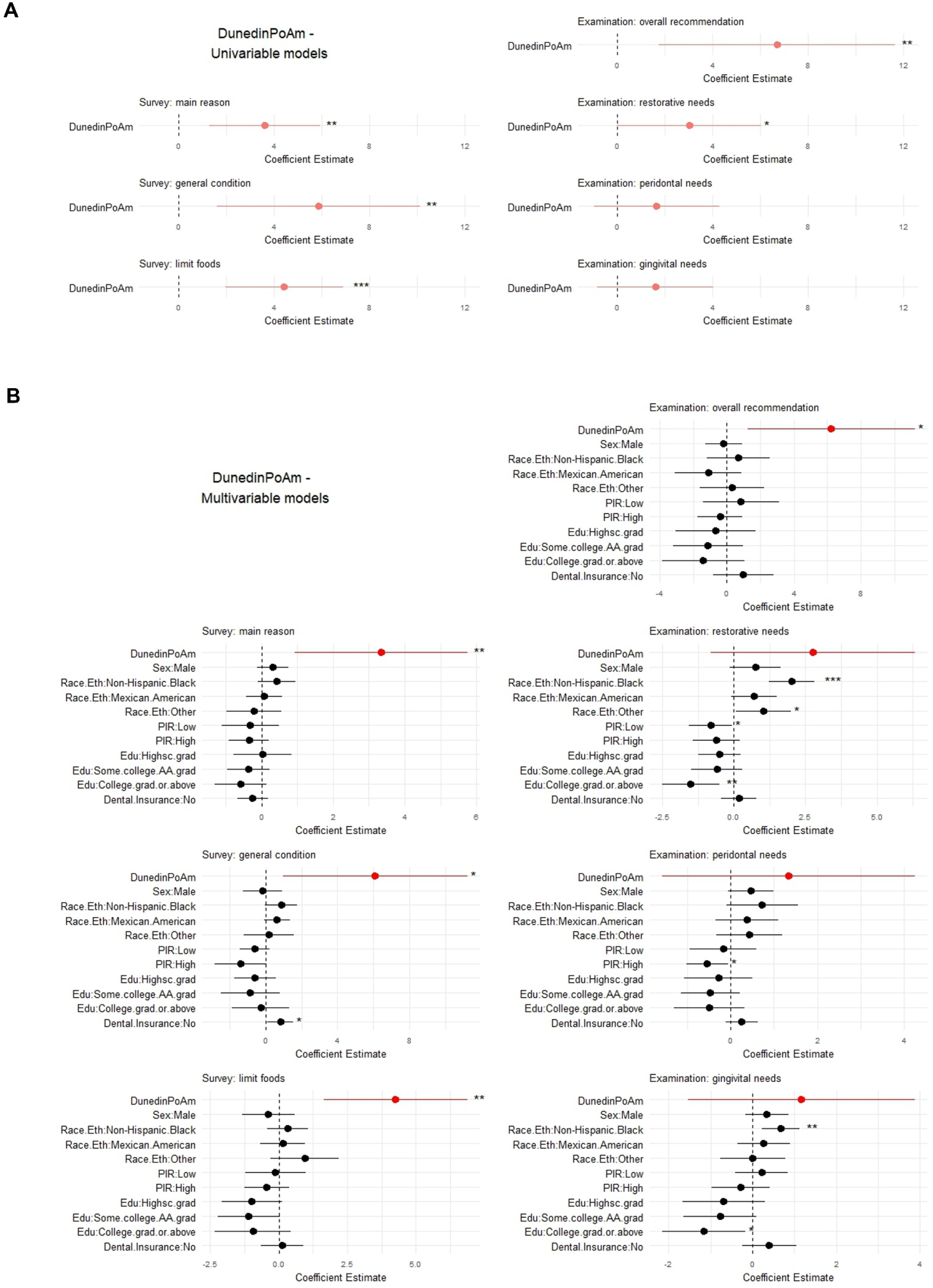
DunedinPoAm is positively associated with various oral health surveys and examination outcomes. Coefficient estimate [95% confidence interval] for the effect of DunedinPoAm on oral health outcomes, encompassing survey (1. whether the main reason for the last dentist appointment was due to an existing oral issue or not for any existing issue; 2. general oral condition self-reported as being poor versus very good, good, or fair; and 3. reported limiting food intake due to teeth problems as always, very often, often, or sometimes versus seldom or never), and examination results (1. overall recommendations after examination were to see a dentist within two weeks versus seeing one at the earliest convenience or continuing with regular routine care; 2. examination reporting untreated caries or restorative needs; 3. examination reporting periodontal needs; and 4. examination reporting gingival needs) are shown in red for **(A)** univariable and **(B)** multivariable models adjusting for sex (reference: female), race-ethnicity (reference: Non-Hispanic White), education (reference: not high school graduate), PIR category (reference: middle), partial or full insurance dental coverage (reference: yes). Stars indicate *p<0.05, **p<0.01, ***p<0.001.

Multivariable models also showed that the last dental appointment being for an existing issue is associated with EAA GrimAge (p=0.0203) and EAA GrimAge2 (p=0.0160), but not others. Poor general condition was associated with EAA Hannum (p=0.0227), EAA PhenoAge (p=0.0377), EAA GrimAge (p=0.0075), and EAA GrimAge2 (p=0.0116). On the other hand, none of the DNAm ages, other than DunedinPoAm, were associated with limiting food due to teeth problems. Apart from DunedinPoAm, only EAA PhenoAge was associated with a recommendation to see a dentist within two weeks (p=0.0191). EAA PhenoAge was also associated with restorative needs (p=0.0405). Collectively, these results suggest that accelerated pace of life (increased DunedinPoAm) and phenotypic or mortality age predictors (EAA PhenoAge, EAA GrimAge, and EAA GrimAge2), may be better indicators of oral health outcomes, compared to chronological age or EAAs based on predicting chronological age.

## Discussion

Previous NHANES studies have only examined the impact of biological age models based on clinical measures on oral health outcomes, while our study comprehensively assessed DunedinPoAm and EAAs of Horvath, Hannum, Weidner, Lin, VidalBralo, PhenoAge, GrimAge, and GrimAge2 epigenetic. Our initial hypothesis was that DunedinPoAm may be most likely to be associated with oral health outcomes compared to other aging clock models because it was developed considering gum health as one of the markers for determining the pace of life. We comprehensively analyzed three survey results and four examination results related to oral health outcomes. Overall. DunedinPoAm was most consistently positively associated with all the survey results and overall examination recommendation to see a dentist within two weeks, even after controlling for sex, race, education, PIR categories, and dental insurance coverage. On the other hand, it was not significantly associated with examination results reporting untreated caries/restorative needs, periodontal needs, and gingival/oral hygiene instruction needs. Therefore, the precise reasons for the overall examination results urging participants to see a dentist in two weeks remain elusive. Self-reported survey outcomes included asking participants whether their last dental appointment was due to an existing oral health reason, and follow-up questions about what those reasons were also unavailable.

Chronological age was associated only with the examination’s overall recommendation result, and it is possible that the appearance of older age may contribute to a bias in the examiner’s recommendation. A reason that chronological age was not generally found to be associated with adverse oral health outcomes, despite what might be expected, could be that the NHANES DNAm data only included age 50 to <85 year olds, thus precluding comparisons across younger and older age, highlighting a limitation in this dataset. The magnitude of the effect for DunedinPoAm was relatively small, which may also be due to the limited age range. Future studies analyzing epigenetic age across a wide range of ages, including below 50 and above 85, are needed. The overall demographic and socioeconomic distributions differed significantly between the below-50 and the 50-or-above groups. The older group showed a relatively larger proportion of female, Non-Hispanic White, high PIR, and lower levels of education. Further studies characterizing demographic and socioeconomic factors across the U.S. by age group are expected to be informative.

Whereas DunedinPoAm was most consistently associated with oral health outcomes, as anticipated, followed by GrimAge, and GrimAge2, then PhenoAge, none of the EAA of aging clock models based on chronological age prediction (Horvath, Hannum, Weidner, Lin, and VidalBralo) were associated with any of the oral health outcomes, suggesting that clocks based on both DNAm and clinical markers tend to be more closely related to oral health outcomes. EAA GrimAge and EAA GrimAge2 were most strongly correlated with DunedinPoAm, followed by PhenoAge, whereas the other EAA models showed marginal or non-significant correlations with DunedinPoAm, consistent with this finding. Future studies identifying specific CpGs that best predict oral health outcomes may elucidate additional underlying molecular mechanisms linking biological aging and oral health. Given that oral health has been linked to a wide range of chronic diseases, characterizing the role of epigenetic age in oral health is expected to be widely significant.

While this study found notable positive associations, their directionality remains elusive due to the use of cross-sectional survey data. A future prospective study would allow analysis of whether accelerated epigenetic aging contributes to or reflects oral health outcomes. Multiple NHANES studies have reported links between periodontitis and biological age measures based on clinical markers,^22–24^ consistent with our results. Among these studies, one further showed accelerated mortality^23^ and another showed increased risk of cognitive outcome,^24^ with accelerated biological age and periodontitis. DunedinPoAm, PhenoAge, GrimAge, and GrimAge2 are all well-characterized as associated with mortality and chronic diseases,^32–35,48,49^ and links between oral health outcomes and mortality and chronic diseases are also well-established.^50,51^ Urinary heavy metal levels was also linked to accelerated epigenetic ages,^17^ and are also known to contribute to oral health outcomes.^52,53^ Future prospective studies may also allow analysis of how adverse oral health outcomes may contribute to chronic diseases and mortality, or whether, in some cases, chronic disease may contribute to worsened oral health, and the potential impact of environmental factors.

While sex was not associated with the oral health outcomes evaluated, specific race-ethnicity groups, and generally, lower education and PIR, were associated with adverse oral health outcomes. Further studies to characterize the relative contributions of demographic and socioeconomic factors to epigenetic processes, versus their direct impact on oral health outcomes, are expected to better inform ways to mitigate adverse oral health outcomes. Finally, DNAm data were only available for 1999-2002 participants, and future studies incorporating recent results would be imperative. Generating DNAm data from samples from more recent NHANES years or a new prospective study would allow for an updated analysis, and commercial consumer kits for different epigenetic ages are widely available and may also be informative if their data were analyzed.

Taken together, analyzing a U.S. population sample, DunedinPoAm was most consistently positively associated with various adverse oral health outcomes, followed by GrimAge and GrimAge2, and PhenoAge, whereas those based on chronological age prediction (Horvath, Hannum, Weidner, Lin, and VidalBralo), were not. Future prospective studies including participants from young to elderly participants widely, are imperative.

## Data Availability

All data produced are available online at the National Health and Nutrition Examination Survey (https://www.cdc.gov/nchs/nhanes/index.html) 1999-2002 cycles.

## Acknowledgments

We gratefully acknowledge the support from the Patricia K. Donahoe Surgeon-Scientist Research Program and Huiying Memorial Foundation to AT.

## Statement of institutional review board waiver

A non-human subjects research determination was obtained from the Mass General Brigham IRB to analyze this data.

## Data Availability Statement

The data used in this study were from the National Health and Nutrition Examination Survey (https://www.cdc.gov/nchs/nhanes/index.html) 1999-2002 cycles.

