## Supplemental Figure S1 for "Specific epigenetic age acceleration measures are associated with oral health outcomes in U.S. adults"

**Supplementary Figure 1**

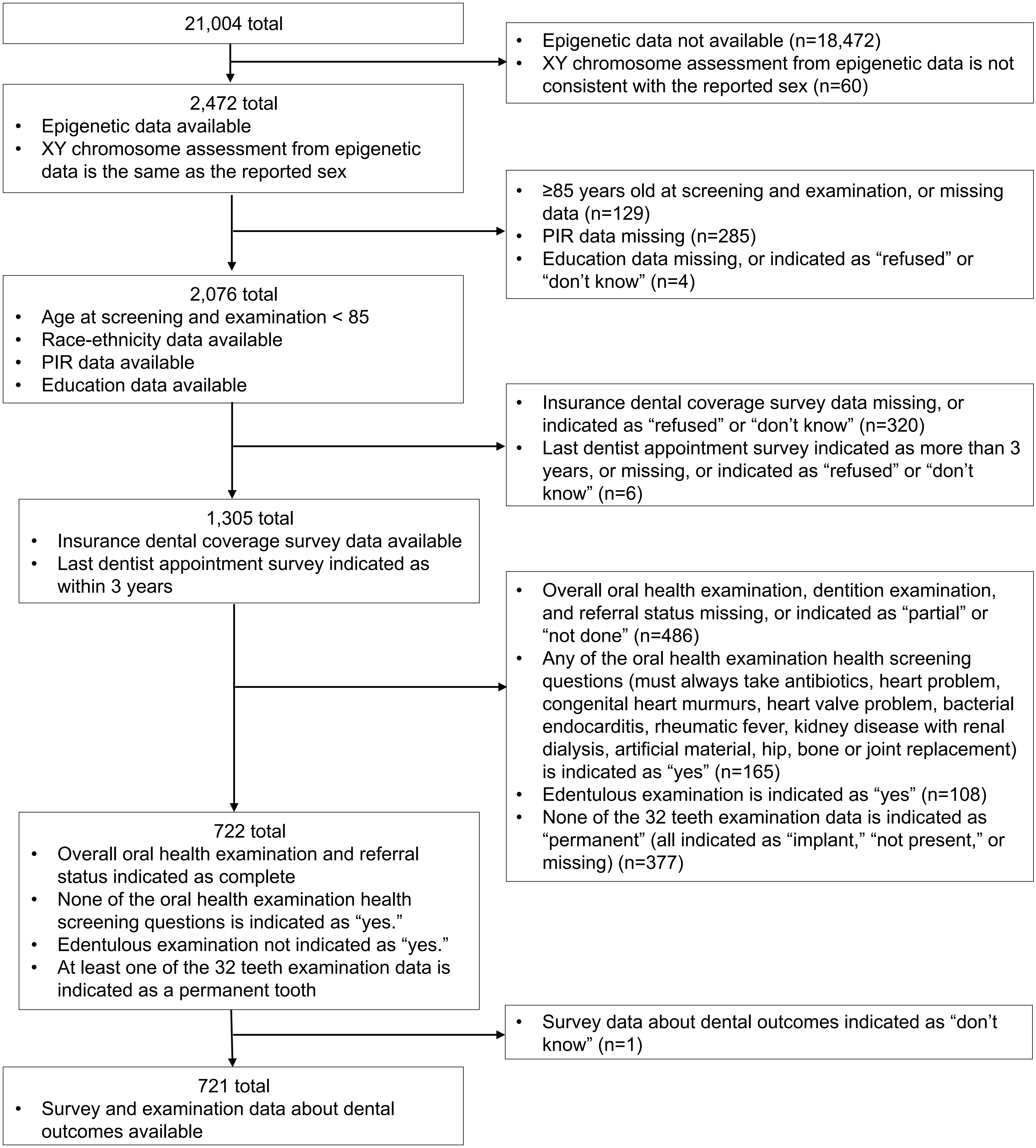


**Supplementary Figure S1.** Flow chart showing the inclusion/exclusion criteria, and the number of participants that were removed at each step and the final number analyzed.
