## Supplemental Figure S2 for "Specific epigenetic age acceleration measures are associated with oral health outcomes in U.S. adults"

### Supplementary Figure 2

## A

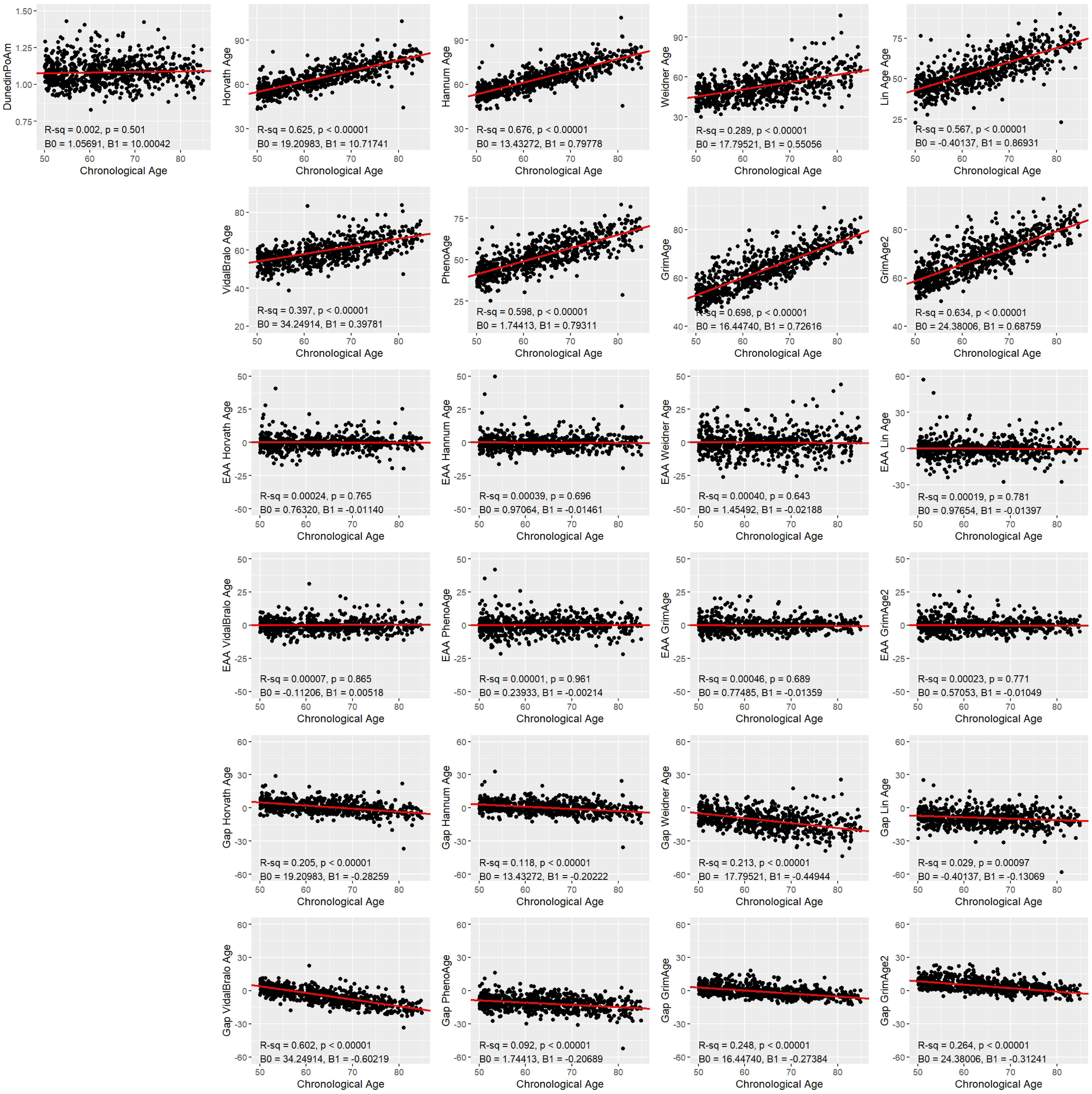

#### Supplementary Figure 2

**B**

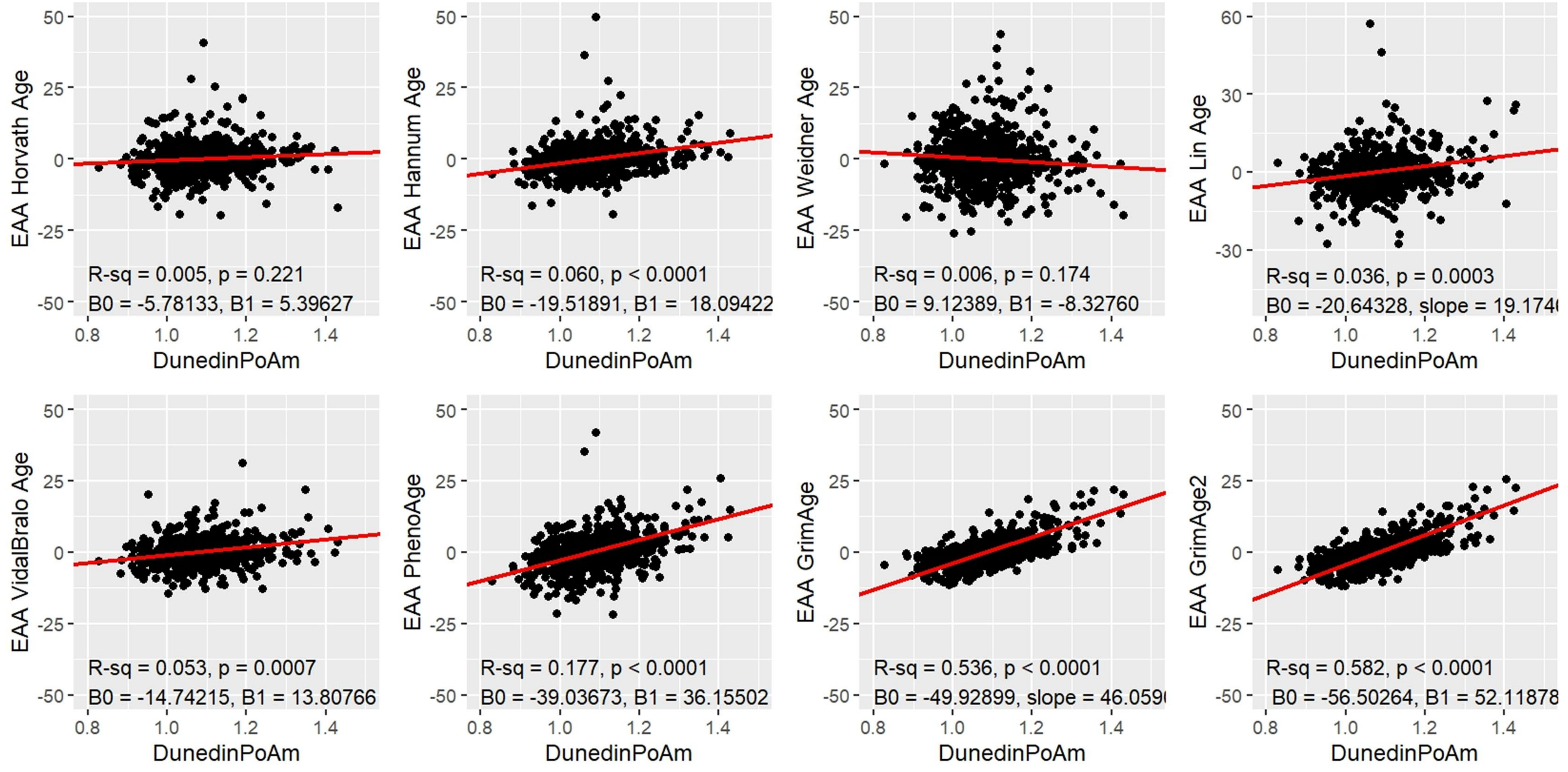


**Supplementary Figure S2.** Linear correlations **(A)** between chronological age and DunedinPoAm, each of the other epigenetic clock models analyzed in this study (Horvath, Hannum, Weidner, Lin, VidalBralo, PhenoAge, GrimAge, and GrimAge2), as well as the epigenetic age acceleration (EAA) for each of them and the gap (*i.e.* difference) between chronological age and each of the epigenetic age, or **(B)** between DunedinPoAm and EAA of the other clocks.
