## Supplemental Table S1 for "Specific epigenetic age acceleration measures are associated with oral health outcomes in U.S. adults"

| **NHANES variable name** | **Outcome – yes** | **Outcome – no** | **Removed** | **Not present in the data corresponding to the other eligibility criteria** |
| --- | --- | --- | --- | --- |
| **Survey results** | | | | |
| OHQ033 – Main reason for last dental visit | 3. Something was wrong, bothering or hurting (n=202);  4. Went for treatment of a condition that dentist discovered at earlier checkup or examination (n=45) | 1. Went in on own for check-up, examination, or cleaning (n=402);  2. Was called in by the dentist for check-up, examination, or cleaning (n=52);  5. Other (n=21) |  | 7. Refused; 9. Don’t know; ∙ missing |
| OHQ010 – General condition of mouth and teeth | 4. Poor (n=71) | 1. Very good (n=190);  2. Good (n=262);  3. Fair (n=199) |  | 7. Refused; 9. Don’t know; ∙ missing |
| OHQ020 - Limit foods because of teeth problems | 1. Always (n=8);  2. Very often (n=11);  3. Often (n=11);  4. Sometimes (n=53) | 5. Seldom (n=75);  6. Never (n=563) | 99. Don’t know (n=1) | 77. Refused; ∙ missing |
| **Examination results** | | | | |
| OHAREC - Overall recommendation for care | 2. See a dentist within the next two weeks (n=19) | 3. See a dentist at your earliest convenience (n=375);  4. Continue your regular routine care (n=328) |  | 1. See a dentist immediately; ∙ missing |
| OHAROCDT - Untreated Caries / Restorative needs | 1. Yes (n=125) | ∙ missing (n=597) |  | 2. No |
| OHAROCGP - Periodontal needs | 1. Yes (n=240) | ∙ missing (n=482) |  | 2. No |
| OHAROCOH - Gingival / oral hygiene instruction needs | 1. Yes (n=237) | ∙ missing (n=485) |  | 2. No |

**Supplementary Table 1.** Outcome definitions used in this study. The original NHANES variable names are provided in the first column, as well as the indications categorized as the outcome in the second column, and indications categorized as no outcome in the third column. Data that were removed from the analysis, or not present in the dataset analyzed, are shown in the fourth and fifth columns, respectively.
