## Supplemental Table S2 for "Specific epigenetic age acceleration measures are associated with oral health outcomes in U.S. adults"

|  | **Overall**  **(N=8,217)** | **Age <50**  **(n=4,563)** | **Age ≥ 50**  **(n=3,654)** | **p-val** |
| --- | --- | --- | --- | --- |
| **Age at examination** | 45.8 ±16.2 | 35.5 ±16.2 | 63.5 ±16.2 | 7.64x10^-36^ |
| **Sex:**  **Female**  **Male** | 4,309 (51.8%)  3,908 (48.2%) | 2,506 (50.8%)  2,057 (49.2%) | 1,851 (53.5%)  1,803 (46.5%) | 0.030 |
| **Race-Ethnicity:**  **Mexican American**  **Non-Hispanic Black**  **Non-Hispanic White**  **Other Hispanic/Other including multi-racial** | 1,958 (7.0%)  1,571 (10.6%)  4,040 (72.1%)  648 (10.3%) | 1,221 (9.0%)  930 (11.8%)  2,017 (68.0%)  395 (11.2%) | 737 (3.5%)  641 (8.5%)  2,023 (79.3%)  253 (8.7%) | 1.13x10^-9^ |
| **PIR category:**  **Low (<1.3)**  **Middle-high (≥1.3 to 4)**  **High (≥4)** | 2,352 (22.0%)  3,579 (41.9%)  2,286 (36.0%) | 1,350 (23.4%)  1,998 (42.5%)  1,215 (34.1%) | 1,002 (19.8%)  1,581 (40.9%)  1,071 (39.3%) | 0.004 |
| **Education:**  **Not high school graduate**  **High school graduate**  **Some college of AA graduate**  **College graduate or above** | 2,708 (20.8%)  1,900 (25.7%)  2,048 (28.8%)  1,561 (24.8%) | 1,275 (17.8%)  1,075 (25.2%)  1,300 (31.5%)  913 (25.4%) | 1,433 (25.9%)  825 (26.4%)  748 (24.1%)  648 (23.7%) | 5.80x10^-5^ |

**Supplementary Table S2.** Using the overall combined 1999-2000 and 2001-2002 demographics data without missing data for age at examination, sex, race-ethnicity, PIR category, and education category (regardless of DNAm or oral health data availability), Survey-weighted mean ± standard deviation for age at examination and unweighted count (weighted %) for other demographic data overall and by chronological age 0 to <50 versus ≥50 years at examination are shown. To compare between the age groups, survey-weighted two-sample t-test p-value for age, or Wald p-values for the other categorical variables are shown.
